# Assessing awareness of blood cancer symptoms and barriers to symptomatic presentation: Measure development and results from a population survey in the UK

**DOI:** 10.1101/2023.04.19.23288810

**Authors:** Laura Boswell, Jenny Harris, Athena Ip, Jessica Russell, Georgia Black, Katriina L Whitaker

## Abstract

**Background:** Low levels of cancer awareness may contribute to delays in seeking medical help and subsequent delays in diagnosis. For blood cancer this may be a particularly prominent problem due to the high prevalence of undifferentiated symptoms such as bodily pain, weakness, nausea and weight loss, resulting in low symptom awareness. The delay is exacerbated by the dismissal of similar symptoms which are often interpreted as mild disease, resulting in multiple consultations prior to diagnosis. This study describes the development of a Cancer Awareness Measure for Blood Cancer (Blood CAM) and presents results from a population-representative survey using the measure.

**Methods:** A rapid systematic review identified constructs relevant to blood cancer. Items were taken from previous awareness measures and other literature and reviewed by expert groups including health care professionals and patients. Cognitive interviews were conducted with ten members of the public to check comprehension and clarity. A total sample of 434 participants completed the survey at Time 1 and n=302 at Time 2 (two weeks later).

**Results:** Internal reliability was high across the different constructs included in the questionnaire (>0.70) and test-retest reliability was moderate to good (0.49-0.79). The most commonly recognised blood cancer symptoms were unexplained weight loss (68.9%) and unexplained bleeding (64.9%) and the least commonly recognised symptoms were night sweats (31.3%) breathlessness and rash/itchy skin (both 44%). In terms of symptom experience, fatigue was the most commonly reported symptom (26.7%) followed by night sweats (25.4%).

Exploratory factor analysis of barriers to presenting at primary care revealed three distinct categories of barriers; emotional, external/practical and service/healthcare professional related. Service and emotional barriers were most common.

**Conclusions:** We developed a valid and reliable tool to assess blood cancer awareness and showed variable awareness of blood cancer symptoms which can help target public health campaigns. We also incorporated additional measures (e.g. confidence to re-consult, ability to understand symptoms) that could be used to tailor public messaging for blood cancer and for other harder to suspect and diagnose cancers.

## Background

The recognition of early diagnosis as a foundation to improve the burden of cancer is unanimous across government and charitable organisations and at global (1) and national levels (2). In the UK, the government has outlined a bold vision for cancer, including that 3 in 4 cancers will be diagnosed at stage 1 or stage 2 by 2028 (3).

Blood Cancer (a group of cancers including leukaemia, lymphoma and multiple myeloma) is the fifth most common cancer in the UK, and the UK’s third leading cause of cancer death (4). It has been hypothesised that low levels of cancer awareness may contribute to delay in seeking medical help and subsequent delays in diagnosis (5). For blood cancer this may be a particularly prominent problem due to the high prevalence of undifferentiated symptoms such as bodily pain, weakness, nausea and weight loss (6, 7), resulting in low symptom awareness (4), and a lack of disease specific knowledge such as night sweats in lymphoma and bleeding and bruising in leukaemia (6). This delay is exacerbated by the dismissal of undifferentiated symptoms which are often interpreted as mild disease, resulting in multiple consultations prior to diagnosis.

In 2007, the Cancer Reform Strategy inspired an action plan for improving earlier diagnosis of cancer, which led, among other initiatives, to the development of a tool to measure public awareness of cancer, the Cancer Awareness Measure (CAM) (8). This measure, which covers classic “alarm” symptoms of cancer (e.g. unexplained lump) is used annually by Cancer Research UK to collect population level data to monitor public awareness over time, compare awareness and attitudes between groups, provide timely targeted evidence for public health campaigns to identify information needs and the measure the impact of these campaigns (9).

Since the original CAM, site-specific versions were developed for ovarian, cervical (10), breast (11), bowel (12) and lung (13) cancers which highlighted important gaps in knowledge, particularly around symptom awareness. However, despite evidence of diagnostic delay in blood cancer patients (14, 15) these diseases have so far received less attention (6).

Our recent rapid systematic review on what causes delays in diagnosing blood cancers (16) found that the majority of studies have focused on the challenges reported by people with blood cancer in how they appraised often vague and non-specific symptoms. There was limited evidence about public awareness of blood cancer symptoms, or about the experiences of patients accessing primary care for their symptoms. We found one study which focused on overcoming primary care related barriers in the diagnostic phase (7). They reported how patients had to advocate for themselves to ensure ongoing investigations and an eventual diagnosis.

Therefore, when considering patient factors in earlier blood cancer diagnosis, it may be important to encompass a broader range of factors/barriers, such as those relevant to patient confidence and peoples’ perceived eligibility for re-accessing healthcare (17). This is also an opportunity to revisit well documented barriers to accessing primary care to see if they can be conceptualised/ considered in ways that will add to the application of findings. For example, previous evidence suggests that people from lower socioeconomic backgrounds may be more likely to report emotional barriers to help-seeking than people from higher socioeconomic backgrounds (5), but there has been no formal exploration of how to delineate emotional versus other barriers.

The aim of this research was to develop and validate a standardised tool to assess blood cancer awareness, and in doing so, include a broader selection of items related to barriers to accessing (and re-accessing) primary care. We addressed the following objectives: a) to develop and validate a blood-specific version of the Cancer Awareness Measure (Blood-CAM), b) to assess the level of blood cancer symptom awareness in the general population, and c) conduct a factor analysis of existing primary care barriers used in Cancer Awareness Measures and describe these, alongside other new constructs (e.g. patient enablement).

## Methods

### Item generation

The tool was conceptualised following a rapid review of literature exploring symptom appraisal, help-seeking and healthcare experiences of people who had had a blood cancer diagnosis (16), as well as drawing on existing tools to measure cancer awareness (10-13) and help-seeking and symptom attribution (18, 19). We sourced blood cancer specific symptoms from Blood Cancer UK (20). The draft scale included nine constructs: symptom awareness (8, 9, 20), re-consultation (21), body vigilance (22), patient enablement (23), social support (24), barriers to help-seeking, symptom experience, attribution and help-seeking (25).

Items were reviewed by experts in blood cancer (N=14) including, oncologists, clinical nurse specialists, nurse advisors, GPs, consultant haematologists and people diagnosed with blood cancer (N=6). The items were reviewed according to four dimensions of interpretability: comprehension, retrieval, decision, and response (26, 27).

Healthcare professional feedback mainly related to the terminology used to describe symptoms which resulted in a few small changes (for example, unexplained pain was separated into bones/ joints and stomach area as these are distinct). Patient feedback most often related to clarity, for example changing the order/ emphasis in sentences (such as “consider the last time you thought about seeking healthcare”).

### Cognitive interviews

The draft tool was used in 10 cognitive interviews with members of the public (age range: 18-66 years) with a range of education backgrounds (6 had GCSEs or lower qualifications) using cognitive probing and think aloud methods (28) to reduce measurement error and assess comprehension and usability of the tool in the target population. Minor amendments were made, for example to respond to feedback related to cultural sensitivity in describing “Paleness (pallor)” this was changed to: “Paleness or unnatural lack of colour/ greying of the skin (pallor)”.

### Online survey

An online survey was carried out to collect data for psychometric analysis. Participants were recruited by a market research company, Dynata (29). Quota sampling was used to ensure the sample was representative of the general population by age, gender and region using 2011 Census and Office for National Statistics (30) data and checked against the England and Wales 2021 Census data (31). Dynata used Quality Score software to exclude participants based on three main data points, 1 ‘Straight lining’, where respondents score the same option from a list of five for every option. 2 ‘Speeders’, participants who complete questions too quickly.3, ‘Passive data and outlier detection’, which monitors in/activity levels e.g. inactive mouse movement.

### Item scoring

For awareness of signs of blood cancer 14 items were scored either 1 (‘Yes, I think this could be a sign of blood cancer’), or 0 (‘No, I don’t think this could be a sign of blood cancer’ and ‘Maybe/ don’t know’), with reverse coding where appropriate (i.e. cough symptom) to indicate whether knowledge was correct or incorrect. Scores were summed to create a total knowledge scale (0-14).

For re-consultation and body vigilance, items were rated on a 5-point Likert from ‘Strongly agree’ to ‘Strongly disagree’ and summed to create a total score (range 3-15).

Each item from the consultancy and body vigilance scales were categorised into endorsed (strongly agree/agree) or not endorsed (neutral to strongly disagree), and the total number endorsed calculated. For patient enablement five items were rated on a four-point response scale of ‘Much better’, ‘Better’, ‘Same’, or ‘Less’ with options for ‘Not applicable’ and ‘I don’t remember/prefer not to say’. Responses were summed to produce a total patient enablement score whereby a higher score indicated greater enablement (score range 5-20). Additionally, these items were categorised into enablement endorsed (much better or better) compared with not endorsed (worse or same).

Social support was assessed from respondents’ answers to four questions taken from the Adult Psychiatric Morbidity Survey, where they were asked to quantify the number of close relationships with adults in their household, with relatives outside their household, with wider friends outside their household plus indicate the number of adults and children living in the household.

Barriers were measured with eighteen items, rated on a Likert scale from ‘Strongly agree’ to ‘Strongly disagree’. Appropriate items were reverse coded and summed whereby higher scores indicated higher barriers (score range 18-90). In addition, each item was categorised into endorsed (strongly agree/agree) vs not endorsed (neutral to strongly disagree), and the total number of barriers endorsed calculated.

For symptom experience, each item was classed as either present (1, ‘Yes’) or not present (0, ‘No’) and the number of potential blood cancer symptoms totalled to produce a symptom experience scale (score range 0-13).

For each potential blood cancer symptom experienced, help-seeking was assessed with the item ‘How long after you first noticed the symptom did you contact your GP practice about it?’ Responses were four incremental time-intervals (up to 2-weeks, between 2 weeks and 1 month, between 1-3 months, and more than 3 months) or did not contact GP. From this, a binary GP help seeking variable was derived for each symptom, No (0) and Yes (1).

Similarly for each symptom present, we assessed cancer symptom attribution with the item about whether they thought cancer could have caused the symptom, yes (1) or no (0).

### Statistical analysis

#### Measurement reliability

For awareness, item difficulty was assessed by calculating the percentage of symptoms correctly identified. The standard difficultly criteria of fewer than 20% (item too difficult) or more than 80% (item too easy) answering correctly was used. Internal reliability (Cronbach’s α and McDonald’s Ω) and item discrimination (item-to-total correlations) were assessed (32). Intraclass correlations (95% CI) were used to assess test-retest agreement over a period of two weeks for the new scales of symptom awareness, re-consultation and barriers to help seeking, using 2-way mixed-effects models (33).

#### Construct validity

To explore the underlying structure of the barrier items and to aid scale reduction we used: exploratory factor analysis using oblique rotation (promax) inspecting scree plots, retaining factor loadings 0.4 or more. Where cumulative variance ≥50%, these were repeated excluding any items where sub-optimal performance was indicated, and an internal reliability of the sub-scales (factors) were computed.

#### Descriptive statistics and logistic regression

Descriptive statistics are presented for sample characteristics (including social support), blood cancer symptom awareness, re-consultation, body vigilance, patient enablement, barriers to help-seeking, symptom experience, attribution and help-seeking.

Multivariable associations between social or demographic factors and likelihood of recognising individual blood cancer symptoms were explored using logistic regression models (as an exploratory analysis, P<0.05 was taken as the level of statistical significance). In these models, we controlled for a priori confounders including age and sex and other covariates that were significant in univariable associations.

### Results

Sample characteristics are shown in Table 1. The sample was broadly representative of the general population according to latest census data (31).

**Table 1.**
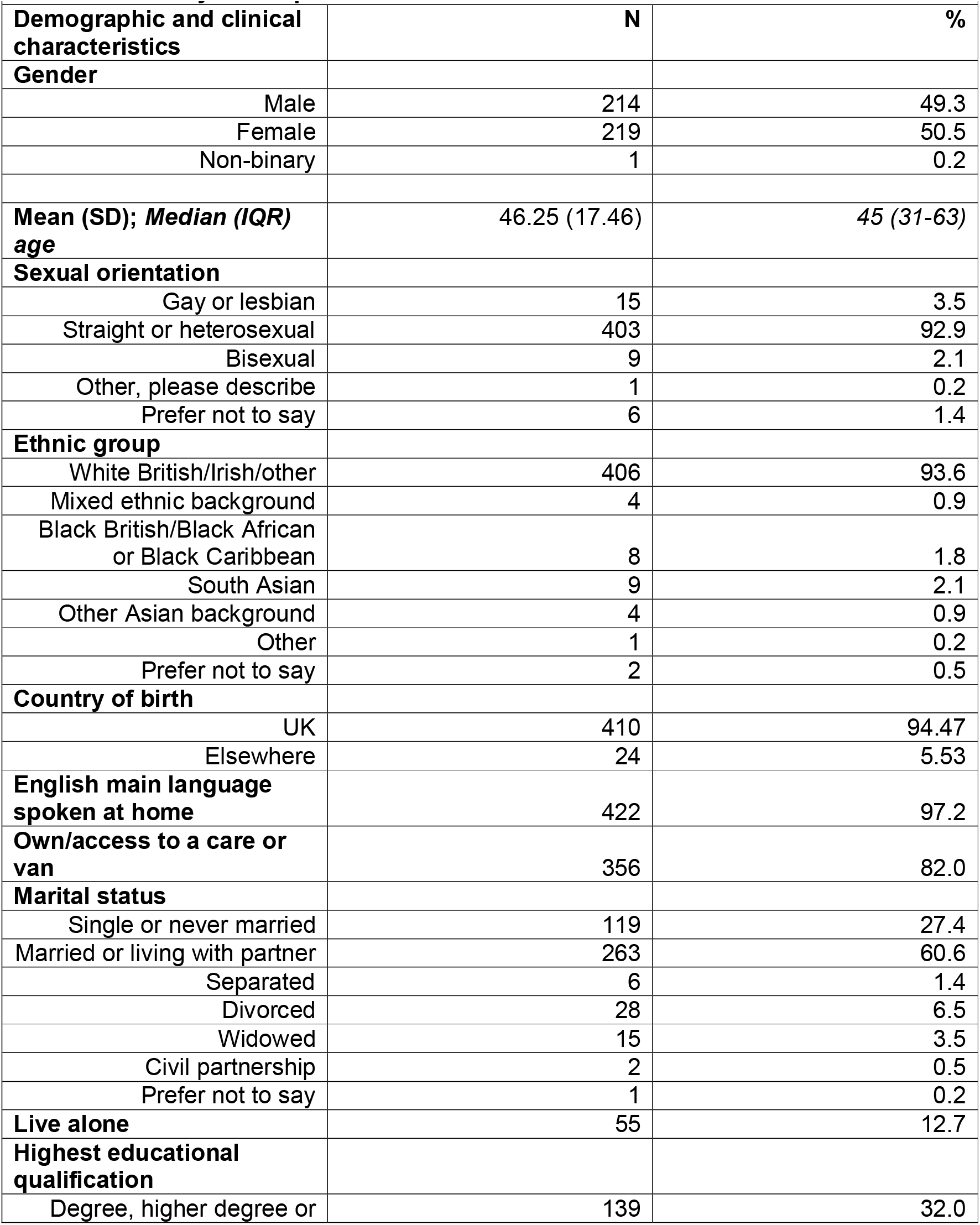
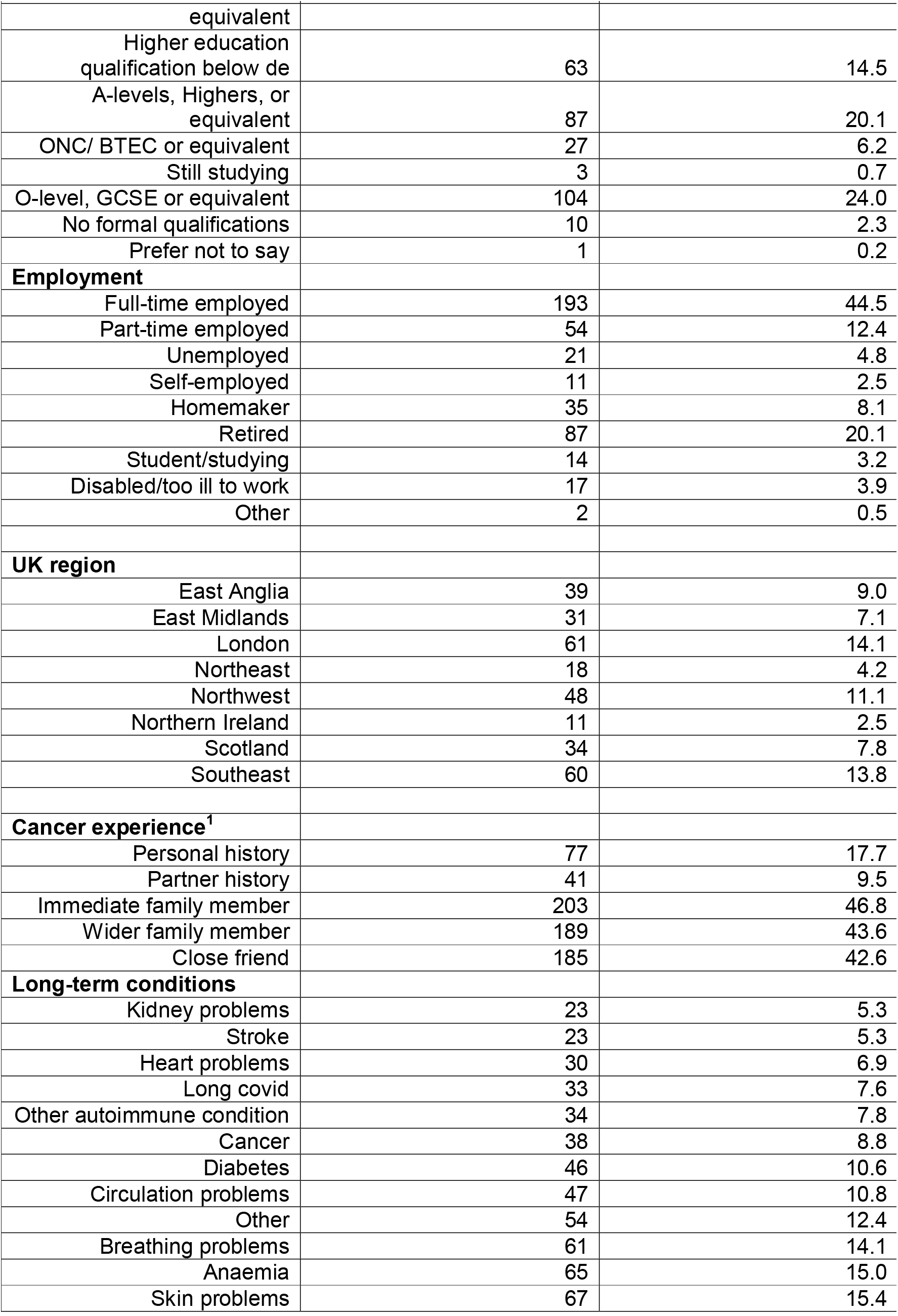
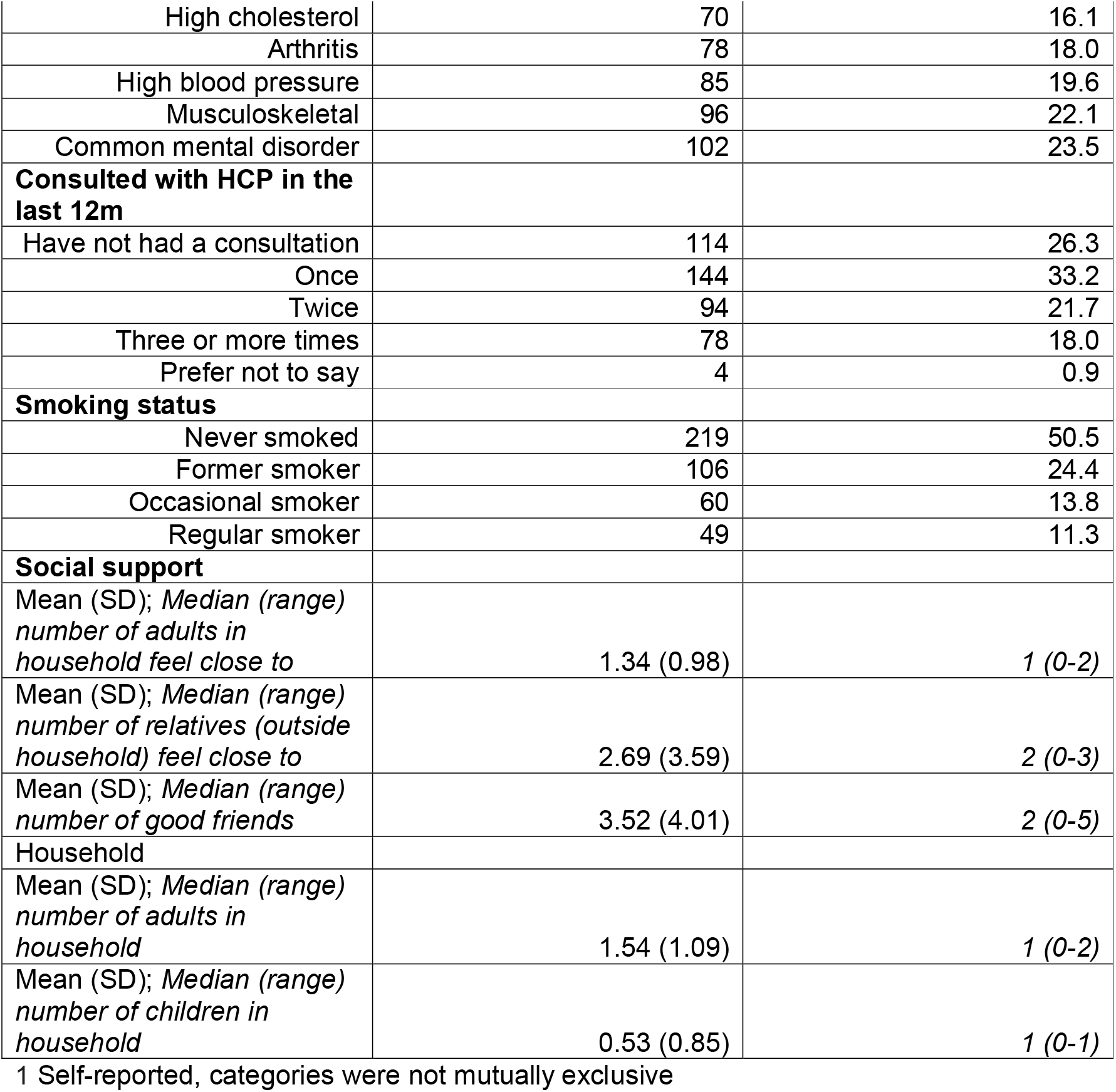
Summary of sample characteristics

#### Symptom awareness

Seven percent (n= 31) of the sample were unable to identify any symptoms. The average number of symptoms recognised was 7 (mean 7.12, SD 4.00; median 7.0, IQR 4-10). The most highly identified symptoms of blood cancer were unexplained weight loss (68.9%), bleeding (64.9%), fatigue (60.6%), bruising (60.1%), lump(s) or swelling (58.5%) and paleness/unnatural pallor (57.1%) (Figure 1). Almost a third of people were aware that cough was not a blood cancer symptom (n=128, 29.5%). More than two-thirds of people were unaware that night sweats were a symptom of breast cancer (n=298, 68.7%).

**Figure 1.**
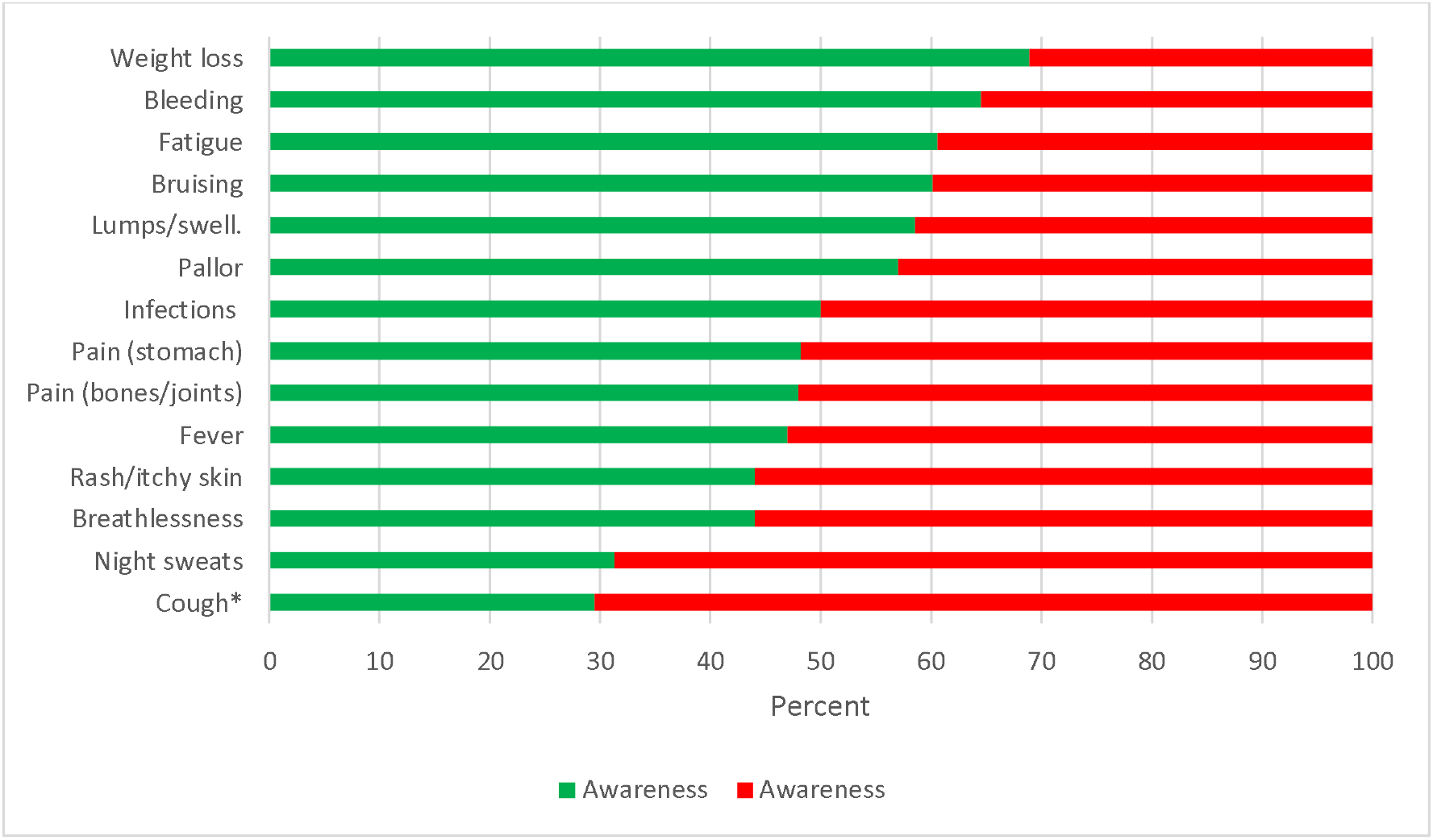
Symptom awareness

All symptom awareness items matched the criterion of being answered correctly by <80% and >20% of participants (Figure 1). Internal reliability of the scale was high (Cronbach’s α 0.85, range 0.83-87, McDonald’s Ω 0.87). Item discrimination supports the ability of individual items to discriminate between those with high or low overall knowledge scores, indicated by item to total correlation ranging from 0.54 to 0.70, with the exception of persistent cough which yielded value of <0.01.Streiner, Norman (32) suggest items with such values <0.02 should be discarded, after which both internal reliability (Cronbach’s α 0.87, range 0.86-87, McDonald’s Ω 0.87) and item discrimination was good (range 0.53-0.71).

Test-retest reliability was good for total number correctly identified (ICC 0.74, 95% CI 0.68-0.79) and moderate to good for individual symptom item (ICC 0.49-0.72) (Appendix Table 1).

Age and gender were consistently associated with being more likely to recognise awareness signs of blood cancer. For example, women were more likely to recognise unexplained bruising, infection, fatigue and pallor as potential signs of blood cancer, compared with men. Older age was significantly associated with being more likely to recognise unexplained bruising, unexplained bleeding, lump/swelling, breathlessness and night sweats. We found the largest gender difference was for unexplained bruising, the odds of recognising this were more than double in women than men (OR=2.27 95%, confidence interval (CI): 1.46–3.54). There were also some significant associations between smoking status and awareness; current smokers were less likely than never smokers to recognise bruising, bleeding, and infection as signs of blood cancer compared with never smokers.

Finally, there were some inconsistent but significant associations between proxies for socioeconomic status and awareness. People who were degree educated were more likely to recognise pain in bones/ joints as a sign of blood cancer compared with those who did not have a degree. However, people who were employed were less likely to identify unexplained bruising as a sign of blood cancer compared with those who were unemployed/economically inactive. People renting were more likely to recognise lump/swelling and pain in bones/ joints than people who owned their own home.

#### Re-consultation, body vigilance and patient enablement

For re-consultations items ‘feeling comfortable about re-consulting if the same symptoms or health problem got worse’ or ‘didn’t get better’ had the highest average scores (mean 4.15, SD 0.85) (Appendix table 2). IQR between 4 and 5 are indicative of a possible ceiling effect although overall the full range of the scale was used by individual participants (range 1-5). Item discrimination was good (all >0.6) as was scale internal reliability (Cronbach’s α 0.79, range 0.66-0.75, McDonald’s Ω 0.79). Body vigilance mean scores were similarly high with being ‘very aware of changes in my body’ having the highest score (mean 4.03, SD 0.80) Both body vigilance and the item ‘I just know when something isn’t right’ narrow IQR scores although again the full scale was used by participants (range 1-5) (Appendix table 2). Item discrimination and internal reliability were all very good (all >0.7 and Cronbach’s α 0.82, range 0.72-0.79, McDonald’s Ω 0.82). Test-retest reliability for the scale was good (ICC 0.61, 95% CI 0.52-0.70) (Appendix table 3). On average, people endorsed two items for both re-consultation and body vigilance (re-consultation mean 2.21, standard deviation 1.04; body vigilance mean 2.26, standard deviation 1.19). People were most likely to say they would re-consult with a doctor symptom ‘didn’t go away or got worse’ with 81.8% (Figure 2).

**Table 2.**
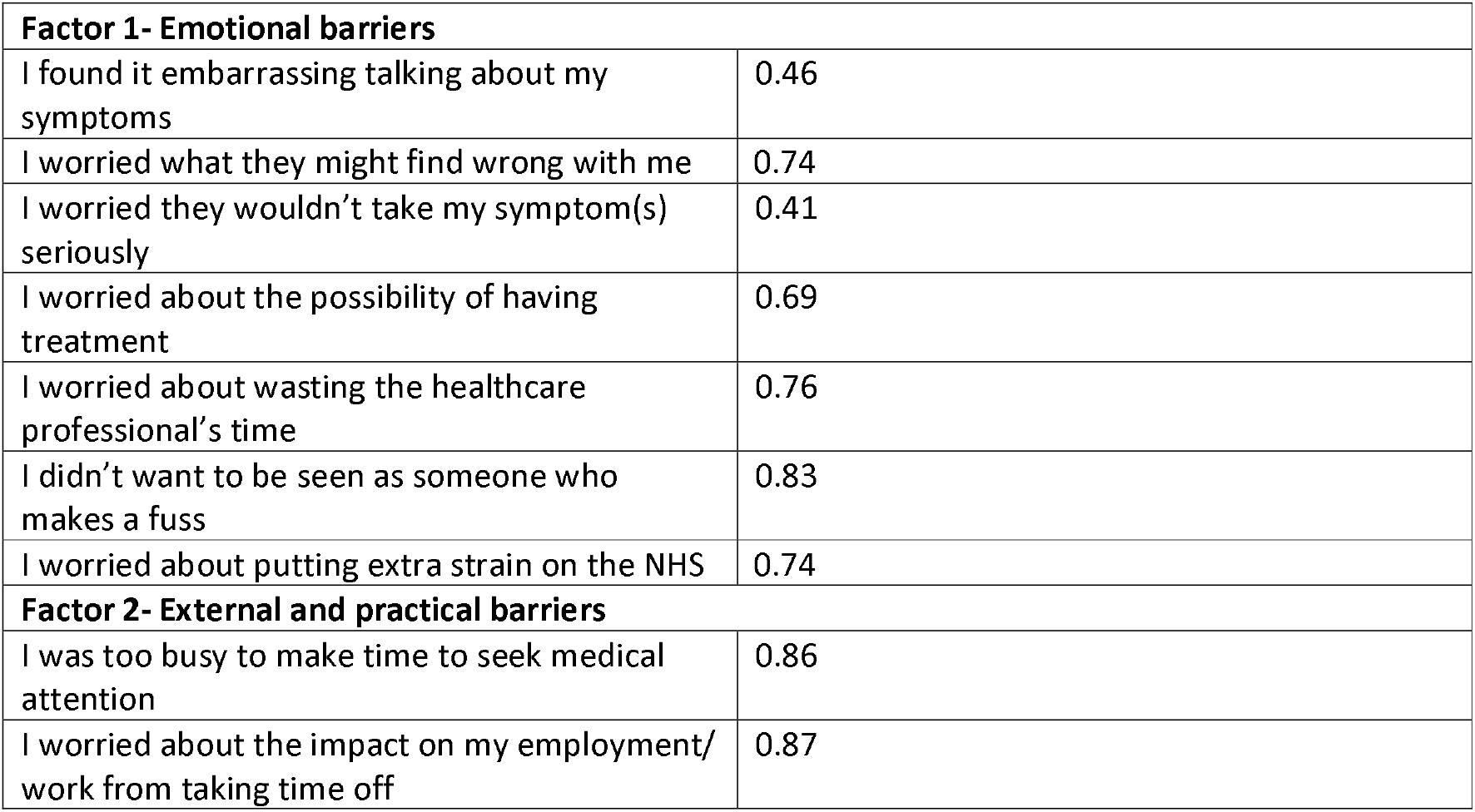
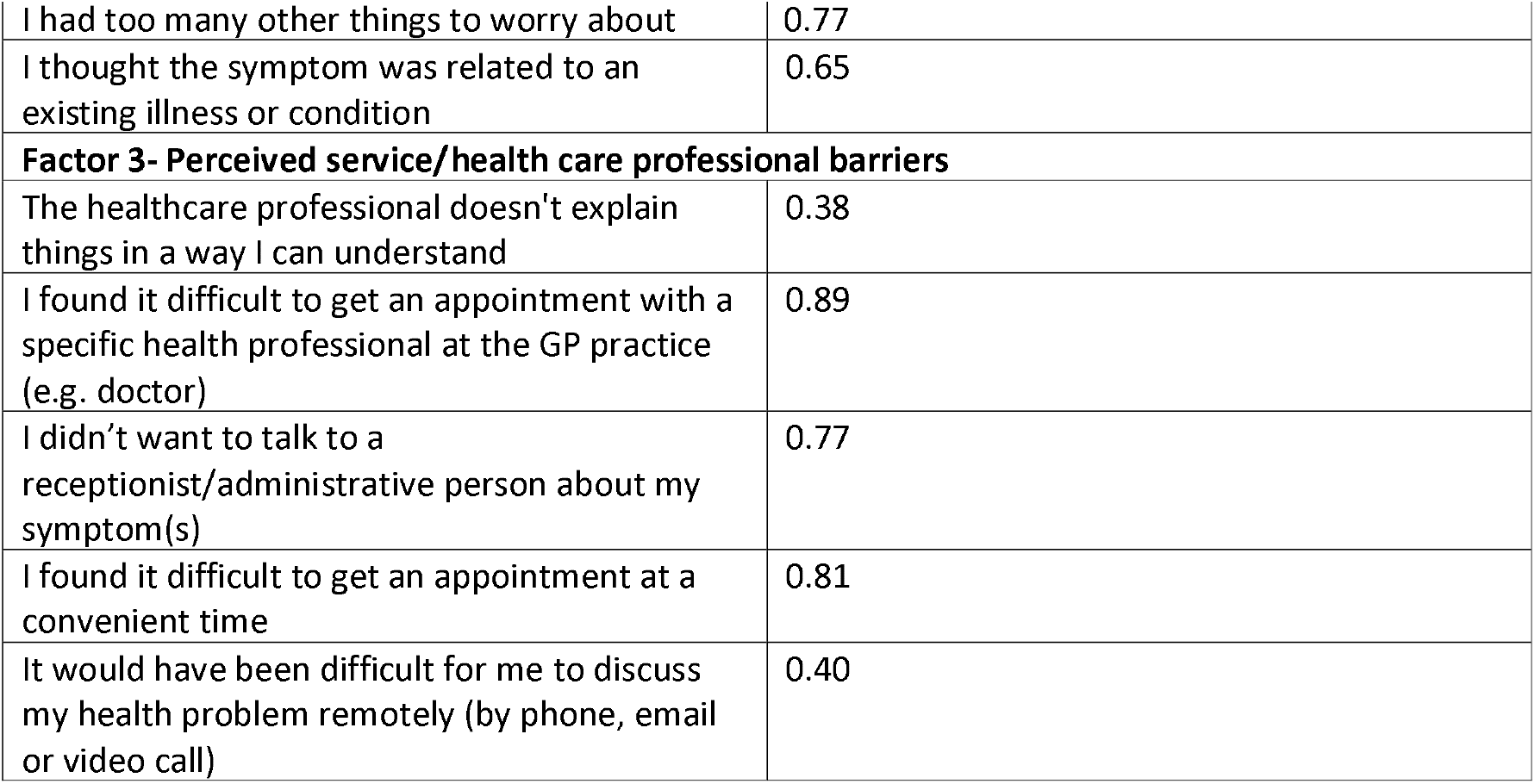
Exploratory factor analysis for the 18 barrier items

| **Factor 1- Emotional barriers** |  |
| --- | --- |
| I found it embarrassing talking about my symptoms | 0.46 |
| I worried what they might find wrong with me | 0.74 |
| I worried they wouldn't take my symptom(s) seriously | 0.41 |
| I worried about the possibility of having treatment | 0.69 |
| I worried about wasting the healthcare professional's time | 0.76 |
| I didn't want to be seen as someone who makes a fuss | 0.83 |
| I worried about putting extra strain on the NHS | 0.74 |
| **Factor 2- External and practical barriers** |  |
| I was too busy to make time to seek medical attention | 0.86 |
| I worried about the impact on my employment/ work from taking time off | 0.87 |
| I had too many other things to worry about | 0.77 |
| I thought the symptom was related to an existing illness or condition | 0.65 |
| **Factor 3- Perceived service/health care professional barriers** |  |
| The healthcare professional doesn't explain things in a way I can understand | 0.38 |
| I found it difficult to get an appointment with a specific health professional at the GP practice (e.g. doctor) | 0.89 |
| I didn't want to talk to a receptionist/administrative person about my symptom(s) | 0.77 |
| I found it difficult to get an appointment at a convenient time | 0.81 |
| It would have been difficult for me to discuss my health problem remotely (by phone, email or video call) | 0.40 |

**Figure 2.**
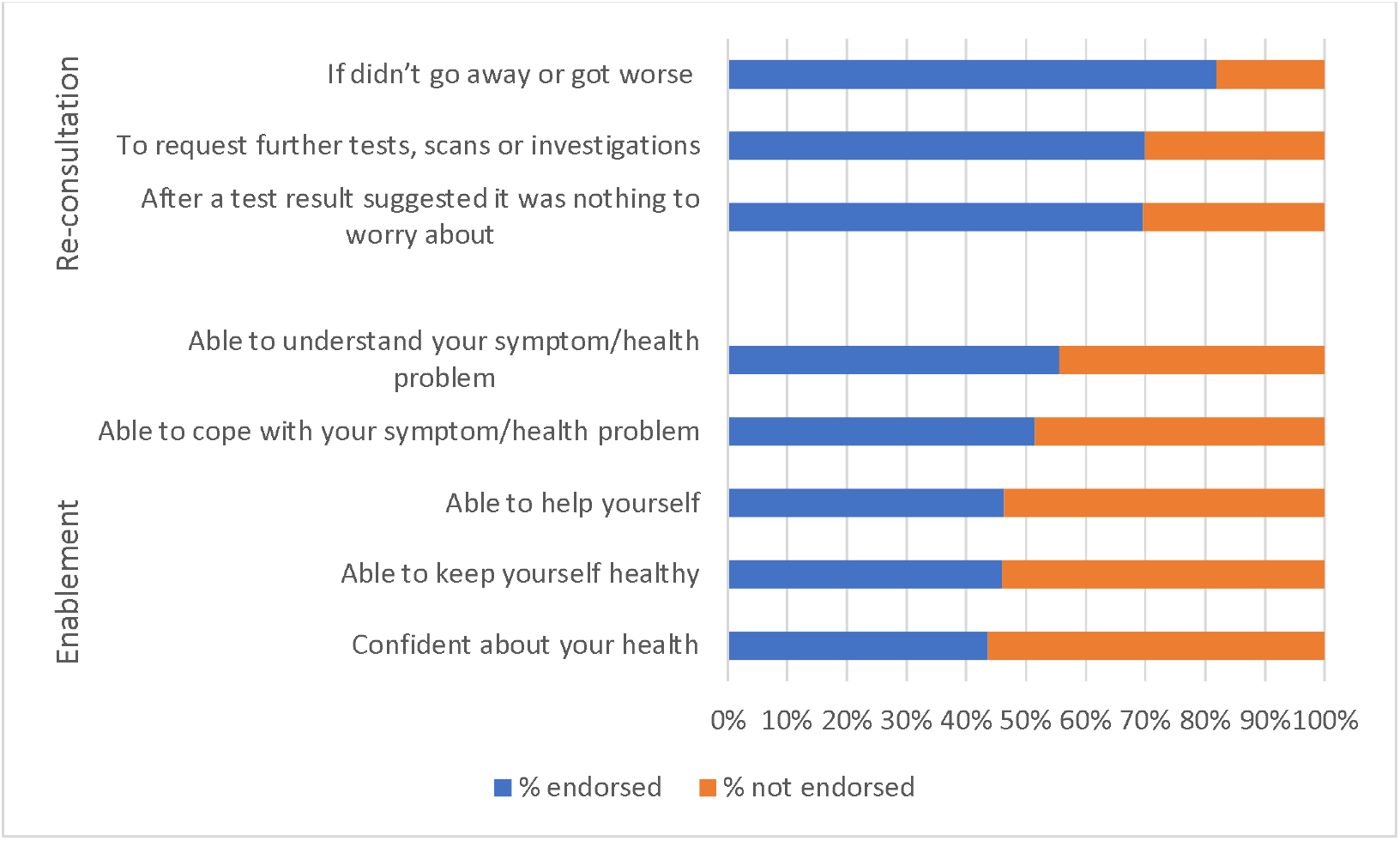
Items endorsed for re-consultation and enablement

Enablement was rated on a scale of 1(Less) to 4 (Much better) with all items mean scores being >2 and ‘able to understand symptom/health problem’ being the item with the highest mean score (mean 2.75, SD 0.85). Again, item discrimination was excellent (all >0.8) as was internal reliability (Cronbach’s α 0.90, range 0.87-0.89, McDonald’s Ω 0.92) (Appendix Table 2). Overall, the mean number of items endorsed as much better or better (“enabled”) was 2.49 (standard deviation 2.12). The most endorsed item for enablement was ‘Able to understand your symptom/health problem’ after visiting a healthcare professional at a GP practice (55.5%) (Figure 2).

#### Barriers to help seeking

The item most likely to be endorsed as a barrier was ‘getting an appointment at a convenient time’ (253, 58.3%) followed by ‘finding it difficult to get an appointment with a specific healthcare professional (HCP) at the GP practice’ (238, 54.8%) and ‘not wanting to be seen as someone who makes a fuss’ (238, 54.8%). ‘I felt I could easily explain/ talk about my symptom(s)’ was the least frequently endorsed (44, 10.1%) (Figure 3). A median of six barriers were endorsed per participant with a mean of 7.2 ranging from 0-17.

**Figure 3.**
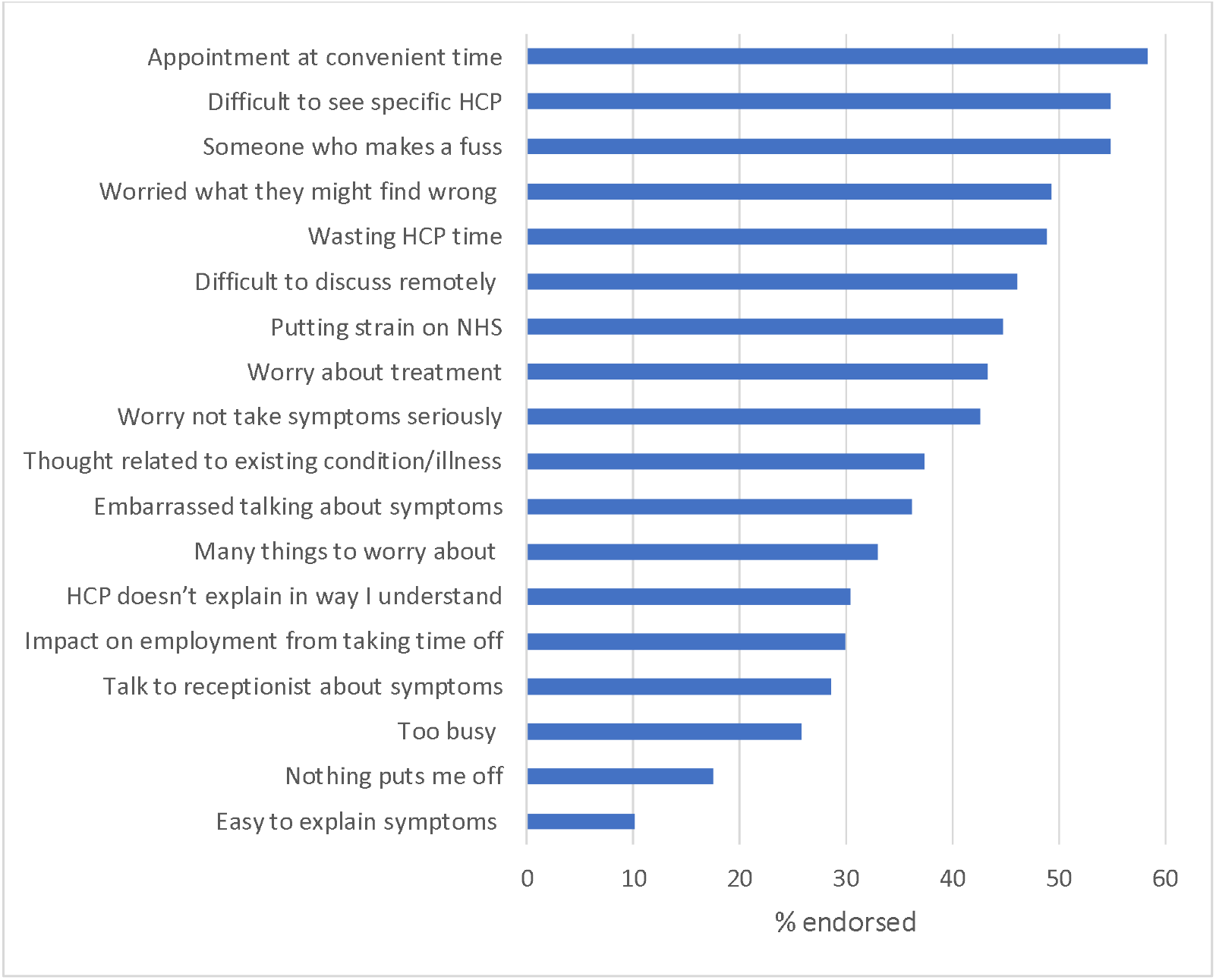
Barriers endorsed

Full descriptive statistics and internal reliability information for the barrier items are shown in Appendix table 4. Internal reliability of the scale was excellent (Cronbach’s α 0.90, range 0.89-91, McDonald’s Ω 0.90). Test re-test reliability was moderate to good (all ICC 0.55-0.84, see Appendix table 5).

### Exploratory factor analysis

The exploratory factor analysis indicated a potential four-factor solution. Further inspection revealed the fourth factor comprised only of the two positive items; ‘Nothing put me off/ delayed me in seeking medical attention’ and ‘I felt I could easily explain/ talk about my symptom(s).’ Examination of the factor correlation matrix indicated that the fourth factor had only weak correlations with the other three factors (<0.1), suggesting that it might not represent the same underlying concept. Subsequently the same analysis was run with these two items removed and a three-factor solution was supported. Consequently, it was decided to retain a three factor solution, additionally supported by weak item-test correlations, internal reliability results for the two items and the possibility that these items could be interpreted as being more closely aligned with self-confidence rather than with barriers.

Therefore, the two items were omitted from the final barriers scale but retained in a different section of the questionnaire. The three final factors represented ‘Emotional barriers’, ‘External and practical barriers’ and ‘Perceived service/health care professional barriers’ (Table 2). Full details of the factor analysis are provided in appendices 6,7 and 8.

### Incidence of symptoms, intervals and attribution

In the last six months, 51.6% (224) had experienced at least one potential blood cancer symptom, with people reporting a mean of two symptoms (SD 2.77, median 1, IQR 0-3). The prevalence of individual symptoms ranged from 8.1% (infections) to 26.7% (fatigue). Overall, 6.7% (29) attributed any potential blood cancer symptom to cancer. Of those with at least one potential blood cancer symptom, half (112/224) had not contacted the GP for any of their reported symptoms over the 6-month timeframe.

## DISCUSSION

We have developed a reliable and valid tool to assess awareness of blood cancer designed to be used in the general population, and for the first time included and assessed the reliability and validity of additional constructs that may be relevant for other cancers where presenting symptoms are vague. For awareness items, internal reliability was high and test-retest reliability was good. For barriers to symptomatic presentation, internal reliability of the scale was excellent and test re-test reliability was moderate to good. We also provided reassurance about the validity/reliability of items added since previous cancer awareness measures, such as those related to body vigilance. Importantly, internal reliability for new/adapted items, for example, candidacy (e.g. confidence in re-consultation) and patient enablement were good or excellent.

Blood cancer awareness was variable; the most recognised blood cancer symptoms were unexplained weight loss and unexplained bleeding, and the least commonly recognised symptoms were night sweats, breathlessness and rash/itchy skin. The level and range of awareness is similar to other site-specific cancer awareness measures, including those measuring lung (13), bowel (12) and ovarian (10) cancer awareness. Symptoms that relate more broadly to cancer (for example those included in the original Cancer Awareness Measure) including unexplained weight loss and unexplained bleeding were among the most recognised and, mirror findings from other site-specific CAMs. This is despite public awareness campaigns usually focusing on specific symptoms associated with different cancer types (e.g. blood in poo for bowel cancer or food sticking when you swallow for oesophageal cancer) (34).

In line with other studies, men had lower knowledge of blood cancer symptoms compared to women (5, 12) which is important given that men are slightly more likely to be diagnosed with blood cancer than women(4). The largest sex difference in this study was for unexplained bruising; the odds of recognising this were more than double in women than men, which could be explained by men engaging less frequently in self-examination (35). We also found that older people had more knowledge of signs and symptoms of blood cancer which is in line with previous research (5, 11, 12) and may be expected given they are at greater risk. Future campaigns to raise awareness of blood cancer may want to target men and younger people.

In addition to exploring blood cancer awareness in the general population, we conducted the first documented factor analysis of barriers to presenting to primary care which revealed three distinct categories; emotional (e.g not wanting to make a fuss), external/practical (e.g. being too busy) and service/healthcare professional barriers (e.g. challenges with making an appointment). The most commonly endorsed barriers were service related (e.g. finding it difficult to get an appointment at a convenient time or getting an appointment with a specific HCP). Emotional barriers, including not wanting to make a fuss and not wanting to waste the doctors time were also commonly endorsed.

This is comparable to previous research which categorised barriers into different groupings (but without conducting a factor analysis). Early work using the original CAM reported that difficulty making an appointment and worry about wasting the doctor’s time were most commonly endorsed (Robb et al., 2009) and this has been a stable finding since (e.g (36) and more recently during the COVID-19 pandemic (19). This means that public awareness campaigns have done little to reassure patients about these barriers. A previous study (37) found that campaigns specifically targeting barriers such as being worried about wasting the doctor’s time did not have an impact and this work should be revisited in future campaigns to understand why these barriers are so prominent and persistent.

Re-consultation, patient enablement and social support were novel constructs included within this CAM and future work can explore their impact on help-seeking outside the scope of the current paper. People generally reported feeling comfortable going back to their GP with the same symptom/ health problem, especially if it didn’t go away or got worse, and slightly less so if it was about following up on further tests or investigations or if a previous test result suggested there was nothing to worry about. Patient enablement was lower, with only half of people reporting that they felt better able to understand their symptom/health problem after a consultation at their GP practice.

Finally, the tool includes measures of symptom experience and help-seeking for potential blood cancer symptoms. Fatigue and night sweats were the most reported symptoms, with over a quarter of the sample experiencing these in the last six months. Of those reporting at least one potential blood cancer symptom, half had not contacted the GP for any of their reported symptoms over the 6-month timeframe and most had not attributed their symptom to blood cancer. This is consistent, if not slightly worse than a recent study looking at help-seeking for general cancer symptoms during the COVID-19 pandemic, which found 45% of people did not contact their GP (19). We will explore this further in another study given it is outside the scope of the aims of this paper.

### Strengths and limitations

A strength of this tool is that it drew on constructs from existing tools to measure cancer awareness (10-13) and included constructs related to re-consultation, patient enablement and social support, which are unique to this study. Previous CAMs and site-specific CAMs were validated between 2009-2012 and therefore the current study provides up to date evidence and reassurance about the ongoing use of CAMs to understand awareness, and barriers to help-seeking in the general population. We have also conducted the first factor analysis of help-seeking barriers which is likely to be useful for future research exploring nuances (e.g. socioeconomic differences in barriers (5, 38)).

A key strength is that we included healthcare professionals, patients and members of the public at all stages of the tool development to ensure that it is applicable to the target population. We also followed recommendations for reporting instrument development (39) and the use of cognitive interviewing means we can be confident about the content validity of the scale.

There were some limitations in this study. The measure was designed to be relevant for all blood cancer symptoms/types combined as it would be too complex to provide separate awareness measures for different types of blood cancer. However, using our tool it would still be possible to explore the awareness of symptoms relevant to specific types of blood cancer. It is not possible to determine the response rate in this study due to the use of participant panels through a market research company, rather than random sampling techniques, however a strength of this approach is that quota sampling was used to ensure the sample was broadly representative of the UK general population. Next steps will be to explore experience and help-seeking for blood cancer symptoms. A further limitation was that much of the analysis was exploratory and larger studies are needed to explore/understand the complex associations with factors such as ethnicity and socio-economic differences.

## Conclusion

This study demonstrates the validity and reliability of a new measure to assess blood cancer awareness in addition to developing useful new constructs that may have applicability beyond blood cancer. The survey revealed lower awareness of blood cancer symptoms amongst men and younger people emphasising the importance for future campaigns to target this group.

## Data Availability

The datasets used and/or analysed during the current study available from the corresponding author on reasonable request.

## Abbreviations

CAM: Cancer Awareness Measure
HCP: Healthcare professional

## Declarations

### Ethics approval and consent to participate

Ethics approval was received from the University of Surrey Ethics committee (FHMS 20-21 024 EGA) and participants provided informed consent. All methods were carried out in accordance with relevant guidelines and regulations.

### Consent for publication

Not applicable.

### Authors’ contributions

GB acquired funding for the study. KLW and GB were responsible for the conceptualization of the study. KLW was responsible for data curation and JH carried out the formal analysis. All authors (LB, JH, AI, JR, GB and KLW) contributed to writing the original draft, reviewing, and editing the manuscript.

## Acknowledgements

Georgia Black is supported by The Health Foundation’s grant to the University of Cambridge for The Healthcare Improvement Studies Institute and this study has been funded by Blood Cancer UK.

## Appendices

**Table 1:** Test re-test for blood cancer awareness

| Signs or symptoms | Correctly identified T1<br>% (N) | Correctly identified T2<br>% (N) | ICC | 95%CI |
| --- | --- | --- | --- | --- |
| Weight loss | 67.55 (204) | 72.52 (72.52) | 0.58 | 0.48-0.67 |
| Bruising | 64.9 (196) | 69.54 (69.54) | 0.72 | 0.65-0.78 |
| Bleeding | 66.89 (202) | 72.19 (218) | 0.60 | 0.49-0.68 |
| Lumps/swell | 62.91 (190) | 63.58 (192) | 0.51 | 0.39-0.61 |
| Breath | 40.4 (122) | 44.04 (133) | 0.60 | 0.49-0.68 |
| Cough | 28.81 (87) | 22.19 (67) | 0.55 | 0.43-0.64 |
| Night sweats | 29.47 (89) | 36.75 (111) | 0.59 | 0.49-0.68 |
| Infections | 51.32 (155) | 52.98 (160) | 0.58 | 0.48-0.67 |
| Fever | 45.7 (138) | 47.35 (143) | 0.59 | 0.49-0.68 |
| Rash/itchy skin | 54.97 (166) | 54.64 (165) | 0.54 | 0.42-0.63 |
| Pain (bones/joints) | 48.68 (147) | 50.66 (153) | 0.49 | 0.36-0.59 |
| Pain (stomach) | 51.32 (155) | 50.66 (153) | 0.52 | 0.40-0.62 |
| Fatigue | 61.59 (186) | 64.57 (64.57) | 0.57 | 0.46-0.66 |
| Pallor | 58.94 (178) | 68.21 (206) | 0.62 | 0.52-0.70 |
|  | Mean (SD)<br>Median (IQR) | Mean (SD)<br>Median (IQR) |  |  |
| Total symptom awareness<sup>1</sup> | 7.24 (4.04)<br>8 (4, 13) | 7.61 (4.07)<br>8 (4, 13) | 0.74 | 0.68 - 0.79 |

<sup>1</sup> For sub-set that answered at both timepoints

**Table 2:** Descriptive statistics and internal reliability for re-consultation, body vigilance and enablement

|  |  |  |  | IQR |  |  |  |  |
| --- | --- | --- | --- | --- | --- | --- | --- | --- |
| Items | Mean | SD | Media<br>n | 25% | 75% | Item-test<br>correl<br>ation | Covari<br>ance | Item $\alpha$ |
| **Candidacy<sup>1</sup>** |  |  |  |  |  |  |  |  |
| *Comfortable going back to my GP with the same symptom/health problem...* |  |  |  |  |  |  |  |  |
| If got worse or didn't go away | 4.15 | 0.85 | 4 | 4 | 5 | 0.59 | 0.44 | 0.75 |
| After a test result suggested there was nothing to worry about, but symptoms persisted | 3.80 | 0.83 | 4 | 3 | 4 | 0.62 | 0.41 | 0.71 |
| To request further tests, scans or investigations | 3.88 | 0.88 | 4 | 3 | 5 | 0.67 | 0.35 | 0.66 |
| *Scale* |  |  |  |  |  |  |  |  |
| Cronbach's $\alpha$ | | | | | | | 0.40 | 0.79 |

| Items | Mean | SD | Media<br>n | IQR | | Item-test<br>correl<br>ation | Covari<br>ance | Item $\alpha$ |
| --- | --- | --- | --- | --- | --- | --- | --- | --- |
|  |  |  |  | 25% | 75% |  |  |  |
| McDonald's $\Omega$ | | | | | | | | 0.79 |
| **Body vigilance**<br>*Thinking about how you view changes in your body* |  |  |  |  |  |  |  |  |
| I am very aware of changes in my body | 4.03 | 0.80 | 4 | 4 | 5 | 0.88 | 0.39 | 0.72 |
| I pay close attention to changes in my body | 3.88 | 0.85 | 4 | 3 | 4 | 0.87 | 0.38 | 0.74 |
| I just know when something isn't right | 3.99 | 0.81 | 4 | 4 | 5 | 0.84 | 0.44 | 0.79 |
| **Scale** |  |  |  |  |  |  |  |  |
| A |  |  |  |  |  |  | 0.41 | 0.82 |
| McDonald's $\Omega$ | | | | | | | | 0.82 |
| **Enablement<sup>2</sup>**<br>*Last time visited HCP at GP practice about symptom/health problem felt...* |  |  |  |  |  |  |  |  |
| Able to understand symptom/health problem | 2.75 | 0.85 | 3 | 2 | 3 | 0.82 | 0.47 | 0.89 |
| Able to cope with symptom/health problem | 2.56 | 0.79 | 3 | 2 | 3 | 0.83 | 0.48 | 0.89 |
| Able to keep yourself health | 2.56 | 0.82 | 2 | 2 | 3 | 0.87 | 0.45 | 0.88 |
| Confident about your health | 2.41 | 0.84 | 2 | 2 | 3 | 0.86 | 0.45 | 0.88 |
| Able to help yourself | 2.58 | 0.88 | 2 | 2 | 3 | 0.88 | 0.43 | 0.87 |
| **Scale** |  |  |  |  |  |  |  |  |
| Cronbach's $\alpha$ | | | | | | | 0.46 | 0.90 |
| McDonald's $\Omega$ | | | | | | | | 0.92 |

1 Rated on a 5-point scale from Strongly agree to strongly disagree. Higher scores indicate higher candidacy
2 Rated on a 4-point scale from Less to Much Better. Higher scores indicate higher enablement

**Table 3:** Test re-test for re-consultation

|  | ICC | 95% CI |
| --- | --- | --- |
| *Comfortable going back to my GP with the same symptom/health problem* |  |  |
| If got worse or didn't go away | 0.54 | 0.42-0.63 |
| After a test result suggested there was nothing to worry about, but symptoms persisted | 0.54 | 0.43-0.64 |
| To request further test, scans or investigations | 0.50 | 0.37-0.60 |
| Total | 0.61 | 0.52-0.70 |

**Table 4:**
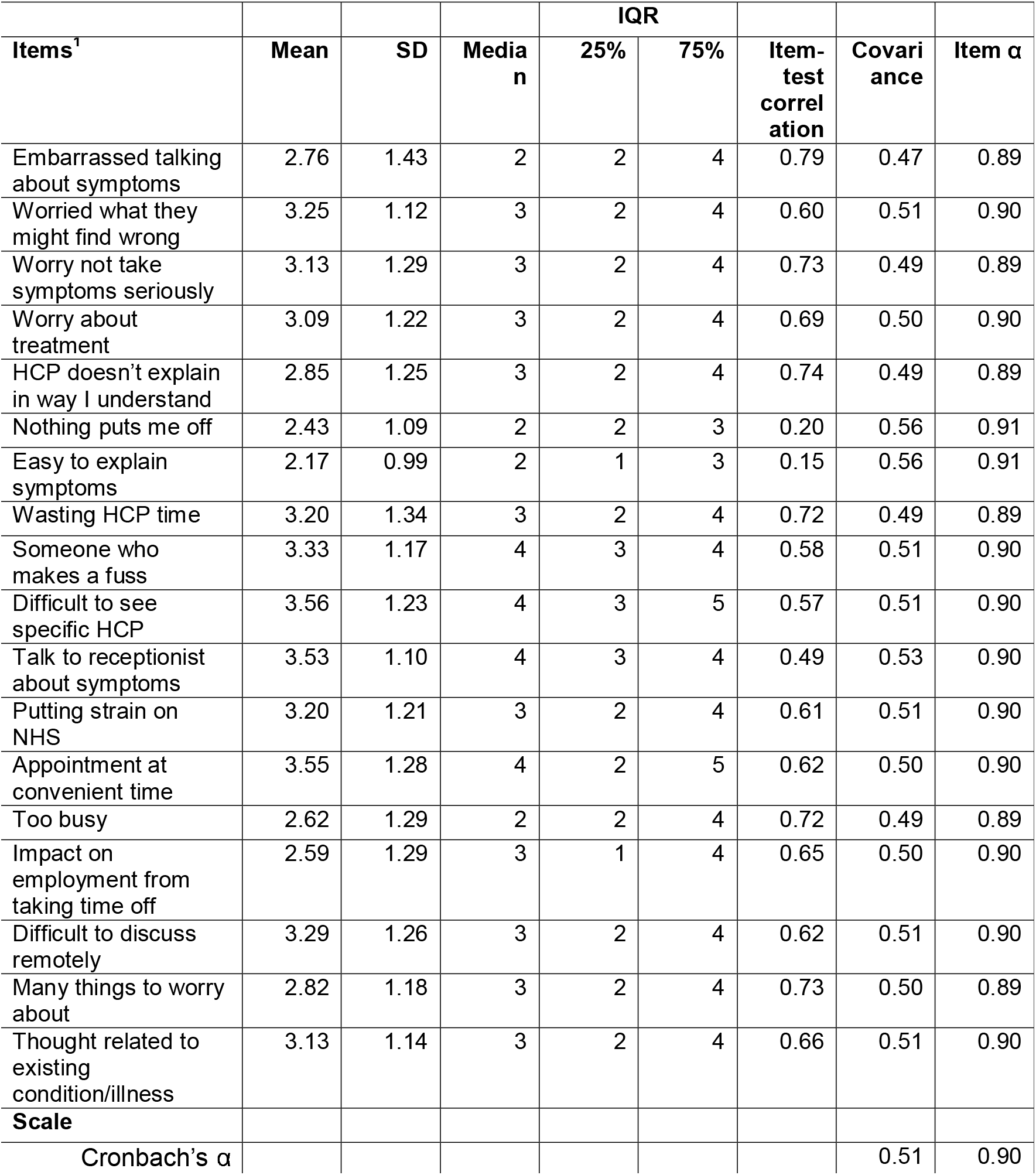
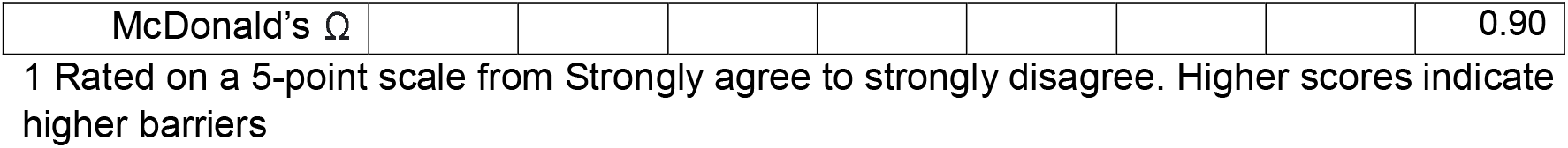
Descriptive statistics and internal reliability of barriers item and scale

| Items<sup>1</sup> | Mean | SD | Median | IQR | | Item-test correlation | Covariance | Item $\alpha$ |
| --- | --- | --- | --- | --- | --- | --- | --- | --- |
|  |  |  |  | 25% | 75% |  |  |  |
| Embarrassed talking about symptoms | 2.76 | 1.43 | 2 | 2 | 4 | 0.79 | 0.47 | 0.89 |
| Worried what they might find wrong | 3.25 | 1.12 | 3 | 2 | 4 | 0.60 | 0.51 | 0.90 |
| Worry not take symptoms seriously | 3.13 | 1.29 | 3 | 2 | 4 | 0.73 | 0.49 | 0.89 |
| Worry about treatment | 3.09 | 1.22 | 3 | 2 | 4 | 0.69 | 0.50 | 0.90 |
| HCP doesn't explain in way I understand | 2.85 | 1.25 | 3 | 2 | 4 | 0.74 | 0.49 | 0.89 |
| Nothing puts me off | 2.43 | 1.09 | 2 | 2 | 3 | 0.20 | 0.56 | 0.91 |
| Easy to explain symptoms | 2.17 | 0.99 | 2 | 1 | 3 | 0.15 | 0.56 | 0.91 |
| Wasting HCP time | 3.20 | 1.34 | 3 | 2 | 4 | 0.72 | 0.49 | 0.89 |
| Someone who makes a fuss | 3.33 | 1.17 | 4 | 3 | 4 | 0.58 | 0.51 | 0.90 |
| Difficult to see specific HCP | 3.56 | 1.23 | 4 | 3 | 5 | 0.57 | 0.51 | 0.90 |
| Talk to receptionist about symptoms | 3.53 | 1.10 | 4 | 3 | 4 | 0.49 | 0.53 | 0.90 |
| Putting strain on NHS | 3.20 | 1.21 | 3 | 2 | 4 | 0.61 | 0.51 | 0.90 |
| Appointment at convenient time | 3.55 | 1.28 | 4 | 2 | 5 | 0.62 | 0.50 | 0.90 |
| Too busy | 2.62 | 1.29 | 2 | 2 | 4 | 0.72 | 0.49 | 0.89 |
| Impact on employment from taking time off | 2.59 | 1.29 | 3 | 1 | 4 | 0.65 | 0.50 | 0.90 |
| Difficult to discuss remotely | 3.29 | 1.26 | 3 | 2 | 4 | 0.62 | 0.51 | 0.90 |
| Many things to worry about | 2.82 | 1.18 | 3 | 2 | 4 | 0.73 | 0.50 | 0.89 |
| Thought related to existing condition/illness | 3.13 | 1.14 | 3 | 2 | 4 | 0.66 | 0.51 | 0.90 |
| **Scale** |  |  |  |  |  |  |  |  |
| Cronbach's $\alpha$ | | | | | | | 0.51 | 0.90 |

|  |  |  |  |  |  |  |  |
| --- | --- | --- | --- | --- | --- | --- | --- |
| McDonald's $\Omega$ | | | | | | | 0.90 |

1 Rated on a 5-point scale from Strongly agree to strongly disagree. Higher scores indicate higher barriers

**Table 5:**
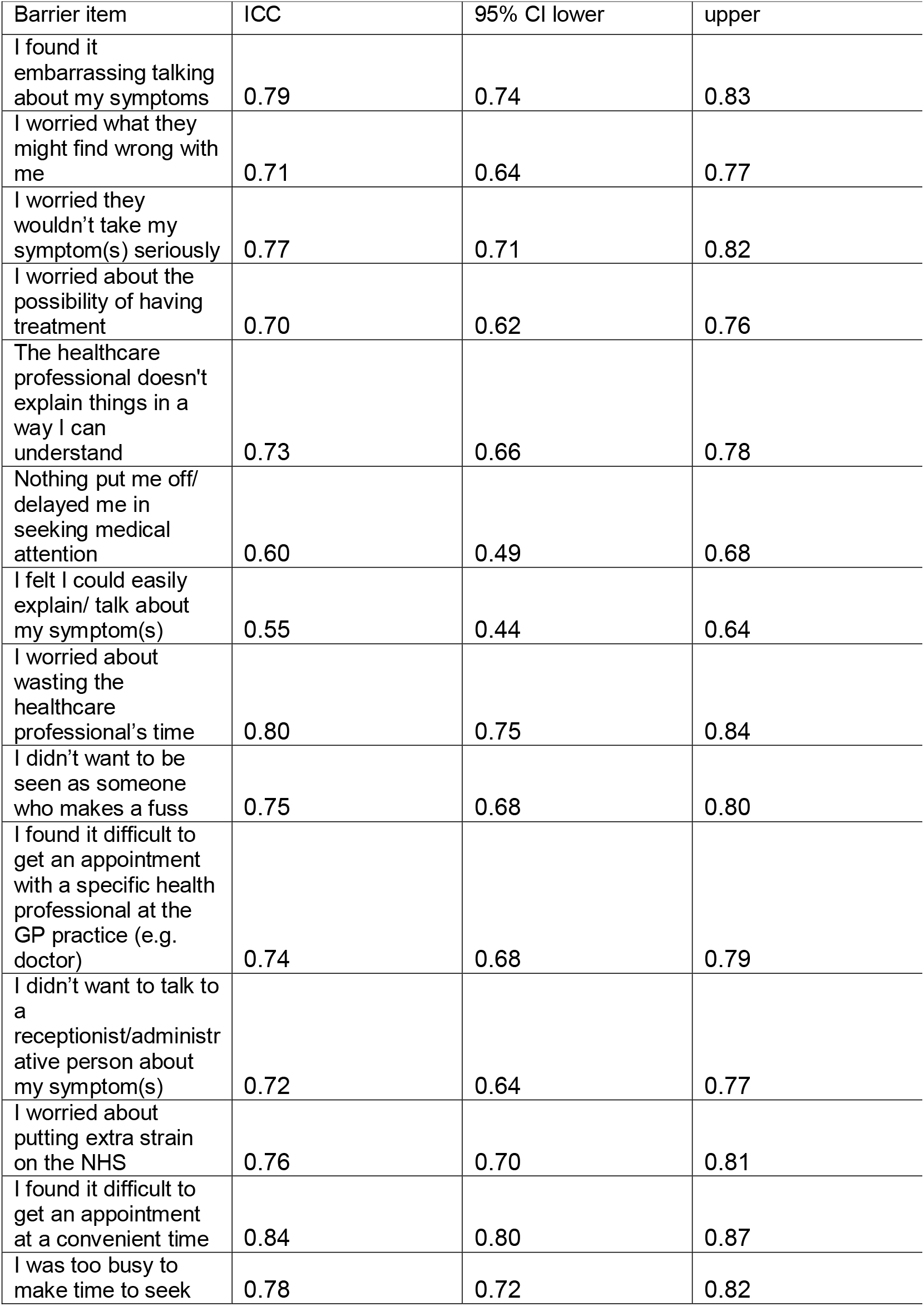
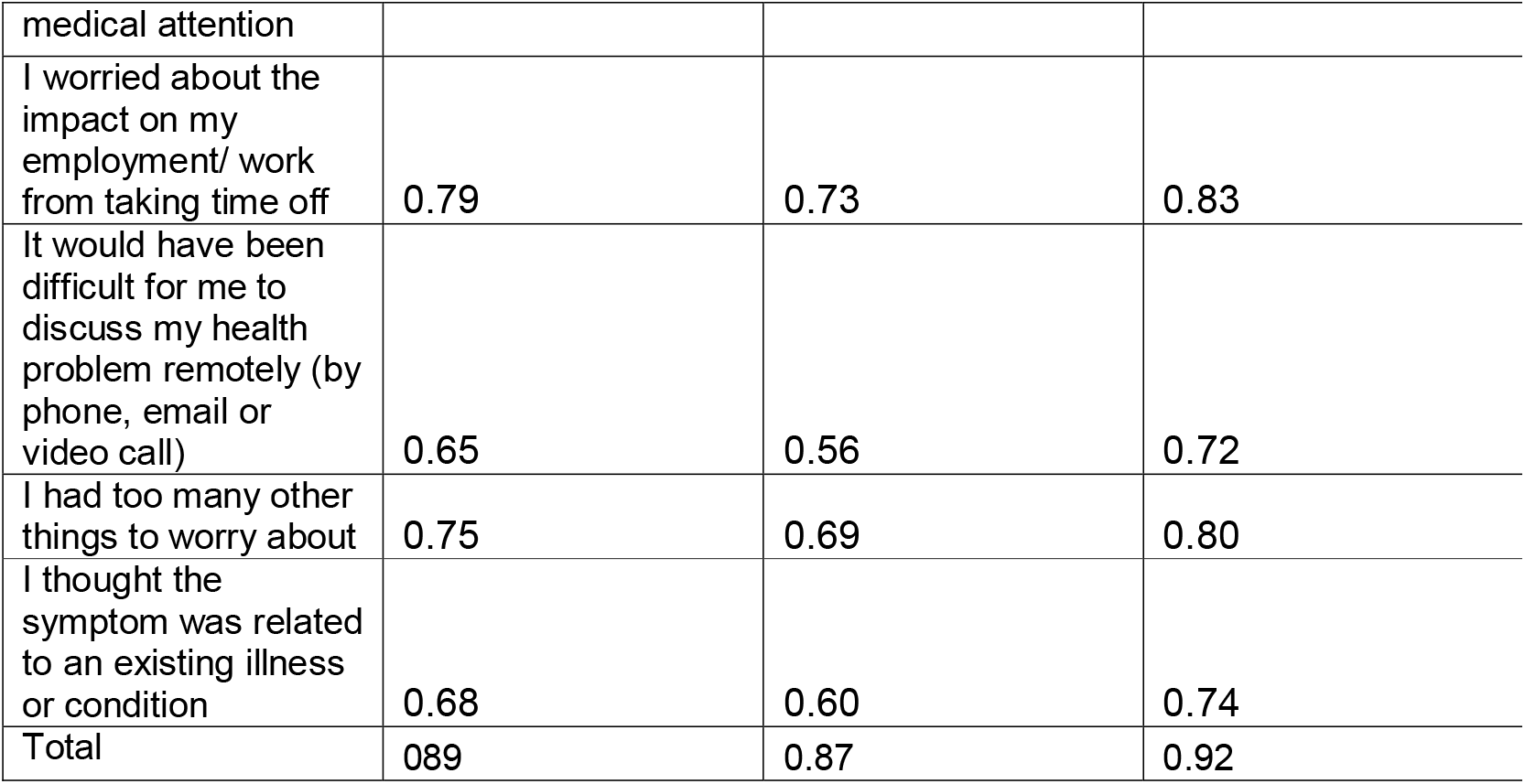
Test re-test for barriers to help-seeking

| Barrier item | ICC | 95% CI lower | upper |
| --- | --- | --- | --- |
| I found it embarrassing talking about my symptoms | 0.79 | 0.74 | 0.83 |
| I worried what they might find wrong with me | 0.71 | 0.64 | 0.77 |
| I worried they wouldn't take my symptom(s) seriously | 0.77 | 0.71 | 0.82 |
| I worried about the possibility of having treatment | 0.70 | 0.62 | 0.76 |
| The healthcare professional doesn't explain things in a way I can understand | 0.73 | 0.66 | 0.78 |
| Nothing put me off/ delayed me in seeking medical attention | 0.60 | 0.49 | 0.68 |
| I felt I could easily explain/ talk about my symptom(s) | 0.55 | 0.44 | 0.64 |
| I worried about wasting the healthcare professional's time | 0.80 | 0.75 | 0.84 |
| I didn't want to be seen as someone who makes a fuss | 0.75 | 0.68 | 0.80 |
| I found it difficult to get an appointment with a specific health professional at the GP practice (e.g. doctor) | 0.74 | 0.68 | 0.79 |
| I didn't want to talk to a receptionist/administrative person about my symptom(s) | 0.72 | 0.64 | 0.77 |
| I worried about putting extra strain on the NHS | 0.76 | 0.70 | 0.81 |
| I found it difficult to get an appointment at a convenient time | 0.84 | 0.80 | 0.87 |
| I was too busy to make time to seek | 0.78 | 0.72 | 0.82 |
| medical attention |  |  |  |
| I worried about the impact on my employment/ work from taking time off | 0.79 | 0.73 | 0.83 |
| It would have been difficult for me to discuss my health problem remotely (by phone, email or video call) | 0.65 | 0.56 | 0.72 |
| I had too many other things to worry about | 0.75 | 0.69 | 0.80 |
| I thought the symptom was related to an existing illness or condition | 0.68 | 0.60 | 0.74 |
| Total | 0.89 | 0.87 | 0.92 |

**Table 6:** Eigenvalues, percentage of variance and cumulative percentage for the four-factor barrier scale

| Factor | Eigenvalue | Percentage of variance | Cumulative percentage of variance |
| --- | --- | --- | --- |
| Factor1 | 7.20 | 40.0 | 40.0 |
| Factor2 | 1.72 | 9.6 | 49.6 |
| Factor3 | 1.56 | 8.7 | 58.3 |
| Factor4 | 1.21 | 6.7 | 65.0 |

**Table 7:** Correlations among four factors

|  | Factor1 | Factor2 | Factor3 | Factor4 |
| --- | --- | --- | --- | --- |
| Factor1 | - |  |  |  |
| Factor2 | 0.52 | - |  |  |
| Factor3 | 0.46 | 0.48 | - |  |
| Factor4 | 0.13 | 0.02 | 0.11 | - |

**Table 8.** Factor loadings and uniqueness for oblique rotation four-factor solution for the 18 barrier items

|  |  |  |  |  |  |
| --- | --- | --- | --- | --- | --- |
| Barrier items | Factor loading |  |  |  | Uniqueness |
|  | Factor 1 | Factor 2 | Factor 3 | Factor 4 |  |
| I found it embarrassing talking about my symptoms | **0.46** | 0.41 | 0.05 | 0.11 | 0.34 |
| I worried what they might find wrong with me | **0.74** | -0.13 | 0.10 | 0.11 | 0.43 |
| I worried they wouldn't take my symptom(s) seriously | **0.41** | 0.11 | 0.37 | 0.14 | 0.41 |
| I worried about the possibility of having treatment | **0.69** | 0.01 | 0.11 | 0.15 | 0.38 |
| I worried about wasting the healthcare professional's time | **0.76** | 0.24 | -0.13 | -0.10 | 0.29 |
| I didn't want to be seen as someone who makes a fuss | **0.83** | 0.01 | -0.17 | -0.05 | 0.41 |
| I worried about putting extra strain on the NHS | **0.74** | 0.11 | -0.07 | -0.23 | 0.40 |
| I was too busy to make time to seek medical attention | 0.07 | **0.86** | -0.05 | -0.02 | 0.24 |
| I worried about the impact on my employment/ work from taking time off | -0.03 | **0.87** | -0.05 | -0.01 | 0.31 |
| I had too many other things to worry about | 0.13 | **0.77** | -0.03 | 0.11 | 0.30 |
| I thought the symptom was related to an existing illness or condition | 0.03 | **0.65** | 0.18 | -0.10 | 0.41 |
| The healthcare professional doesn't explain things in a way I can understand | 0.25 | 0.33 | **0.38** | -0.02 | 0.40 |
| I found it difficult to get an appointment with a specific health professional at the GP practice (e.g. doctor) | -0.11 | 0.01 | **0.89** | -0.06 | 0.28 |
| I didn't want to talk to a receptionist/administrative person about my symptom(s) | 0.23 | -0.33 | **0.77** | -0.06 | 0.41 |
| I found it difficult to get an appointment at a convenient time | -0.21 | 0.23 | **0.81** | 0.02 | 0.28 |
| It would have been difficult for me to discuss my health problem remotely (by phone, email or video call) | 0.01 | 0.39 | **0.40** | -0.03 | 0.53 |
| Nothing put me off/ delayed me in seeking medical attention | -0.02 | 0.04 | -0.05 | **0.87** | 0.25 |
| I felt I could easily explain/ talk about my symptom(s) | -0.03 | -0.02 | -0.03 | **0.88** | 0.23 |

